# Genomic characterization, origin, and local transmission of Oropouche Virus in Bolivia in 2024

**DOI:** 10.1101/2024.12.23.24319382

**Authors:** Joel Alejandro Chuquimia Valdez, Ighor Arantes, Sebastián Sasías Martínez, Cleidy Orellana Mendoza, Nelly Mendoza Loayza, Jhonatan D. Marquina, Helen Castillo Laura, Roxana Salamanca Kacic, Maya Xochitl Espinoza Morales, Lionel Gresh, Mariela Martínez Gómez, Jairo Méndez-Rico, Gonzalo Bello, Felipe Gomes Naveca, Leidy Roxana Loayza Mafayle

## Abstract

**Background:** The Oropouche virus (OROV) is an arthropod-borne virus that causes an acute febrile illness similar to other arboviral diseases. In 2024, Oropouche cases sharply increased in several countries of the Americas, including Bolivia. Here, we performed a genomic study to investigate the origin and spread of OROV in the Bolivian Amazon region.

**Methods:** Full-length OROV genomes from 34 positive samples collected in the three affected Bolivian departments during 2024 were sequenced using an amplicon-based approach. Maximum Likelihood (ML) phylogenetic analyses of separate viral segments were conducted to identify the responsible viral lineage. Bayesian phylogeographic analysis of concatenated viral segments was used to reconstruct the viral spatiotemporal dispersion pattern within the country.

**Findings:** Epidemiological data shows that the first OROV-positive cases in 2024 in Bolivia were reported in samples collected from the Pando department during mid-January, and the peak of OROV-positive cases occurred in mid-April. The phylogenetic analysis of OROV genomes revealed that all cases detected in Bolivia belong to the novel reassortant OROV clade that drove the recent epidemic in Brazil. Our phylogeographic analysis detected at least two exportation events of OROV from the Brazilian state of Acre to the Bolivian municipalities of Guayaramerín and Riberalta, both located in the Beni department, with subsequent dissemination to municipalities of Pando and La Paz departments. Viral introductions probably occurred between early October and early November 2023, thus indicating a lag of about three months between OROV introduction and detection in Bolivia.

**Interpretation:** Our findings confirm that OROV spread at least twice from the western Brazilian Amazon to the neighboring Bolivian department of Beni in late 2023, successfully establishing regional transmission chains. The introduction and spread of OROV in Bolivia coincides with the Amazonian rainy season, from November to April, a period likely marked by an increase in vector abundance. These findings underscore the critical need for active OROV surveillance across the cross-border Amazonian region between Brazil and Bolivia. They also confirm the potential for sustained OROV transmission within the Bolivian Amazon, highlighting the importance of preparedness for future outbreaks.

**Funding:** This publication was in part supported by the Cooperative Agreement Number NU50CK000639 awarded to the Pan American Health Organization and funded by the Centers for Disease Control and Prevention. Its contents are solely the responsibility of the authors and do not necessarily represent the official views of the Centers for Disease Control and Prevention or the Department of Health and Human Services.

**Research in context:** *Evidence before this study:* Before 2024, large outbreaks of Oropouche virus (OROV) were predominantly reported in the Amazon regions of Brazil and Peru. However, in 2024, significant outbreaks first emerged in the Brazilian Amazon region. They were soon followed by a surge of cases in the neighboring South American countries of Bolivia, Colombia, and Peru. We searched PubMed and preprint servers (medRxiv and bioRxiv) available as of October 25, 2024, for studies examining the circulation of OROV in Bolivia, using the terms [“Oropouche” AND “Bolivia”]. We identified only one study that reported a few anecdotal cases of past OROV infections in Bolivia, relying on serological tests and a couple of reviews.

*Added value of this study:* This is the first study to analyze the genomic characteristics of OROV circulating in Bolivia. In this study, we sequenced 34 full-length OROV genomes, representing 10% of all RT-qPCR-confirmed OROV cases across Pando, Beni, and La Paz departments between January and May 2024. The OROV detected in Bolivia belongs to the novel reassortant lineage recently identified in Brazil. We identified at least two introductions of OROV from the western Brazilian Amazon region into the neighboring Bolivian department of Beni around late 2023, followed by its spread to other regions within Bolivia during the rainy season. Our estimates indicate that the virus circulated in Bolivia for approximately three months before the first case was detected.

*Implications of all the available evidence:* Our study confirms that the novel OROV reassortant lineage recently identified in Brazil rapidly disseminated across the Amazonian border into Bolivia. The successful establishment of OROV in Bolivia indicates that the country possesses suitable ecological conditions to support sustained transmissions of this arbovirus. Our findings also emphasize the crucial need for active and sustained molecular surveillance of OROV in the Bolivian Amazon region to enable the timely detection of new outbreaks in the country.

## Introduction

The Oropouche virus (OROV) is an arthropod-borne virus primarily transmitted by the bite of the hematophagous midge *Culicoides paraensis*. The symptomatic infection with this virus causes Oropouche fever, an acute febrile illness similar to other febrile diseases caused by co-circulating arboviruses such as dengue and chikungunya (1). OROV or *Orthobunyavirus oropoucheense* belongs to the Simbu serogroup within the *Orthobunyavirus* genus of the *Peribunyaviridae* family. This virus has a negative-sense, single-stranded RNA genome composed of large (L), medium (M), and small (S) segments (1).

OROV was first detected in Trinidad and Tobago in 1955, and most outbreaks until 2000 were reported in Brazil and Peru (1). Over the past 25 years, sporadic cases of OROV have been identified in other American countries, including Argentina, Bolivia, Colombia, Ecuador, French Guiana, Haiti, and Panama (1). In 2024, the number of OROV cases sharply increased in the Americas, with a total of 10,183 confirmed cases as of the epidemiological week 40 of 2024, including two deaths (2). Autochthonous Oropouche fever cases have been reported across six countries: Brazil (*n* = 8,258), Peru (*n* = 936), Cuba (*n* = 555), Bolivia (*n* = 356), Colombia (*n* = 74), Ecuador (*n* = 2), and Guyana (*n* = 2) (2). The recent expansion of the virus outside the Amazon basin and the broad distribution of its primary vector have raised serious concerns about the potential spread of OROV beyond its historical range (2).

The presence of OROV in Bolivia was first detected in Cochabamba as part of a clinical surveillance program investigating febrile disease etiologies from 2005 to 2007 (3). However, this study analyzed a limited number of locations in Bolivia and only used serological tests for OROV detection. Furthermore, the genetic characteristics and geographic distribution of the OROV circulating in Bolivia remained unclear. Systematic surveillance of OROV was not implemented in Bolivia until 2024, when the Pan American Health Organization (PAHO) issued an epidemiological alert for OROV in the Americas (4). Between January and May 2024, the Ministry of Health and Sports reported a total of 356 positive OROV cases confirmed by RT-qPCR in both rural and urban areas across the departments of Pando, Beni, and La Paz, all located within Bolivia’s Amazon basin (2,5).

In this study, we sequenced 34 OROV-positive samples collected from the three affected departments during 2024. Our aim was to identify the viral lineage responsible for the current Oropouche fever outbreak in Bolivia and to reconstruct the viral spatiotemporal dispersion pattern within the country.

## Material and methods

### OROV positive samples and epidemiological data

Clinical samples from patients with suspected OROV infection were sent to the National Center for Tropical Diseases (CENETROP) of Bolivia from the departments of La Paz, Beni, and Pando for OROV detection. The definition of a suspected case of Oropouche was as follows: any person who resides in or has visited in the last 14 days areas of transmission or with a history of Oropouche outbreak and who presents at least one or more of the following signs and symptoms: fever greater than or equal to 38°C, intense headache, chills, arthralgias, lack of appetite, myalgias, photophobia, dizziness, lumbar pain, difficulty walking. Molecular detection of OROV was performed using the duplex real-time PCR assay developed by Naveca *et al*. (6), which PAHO has recommended for the molecular surveillance of Oropouche and Mayaro viruses (7). From the 156 OROV-positive samples identified by CENETROP between January and June 2024, 34 samples (22%) were selected for whole-genome sequencing based on Ct values (less than 25), epidemiological week (EW), and department of detection. Each selected sample had an associated identification code and associated epidemiological metadata such as age, sex, spatial location, date of symptoms onset, collection date, date of notification, and hospitalization (**Table S1**).

### Whole-genome sequencing and genome assembling

Complete genome sequencing was carried out with an amplicon-based strategy previously developed by the Emergent, Reemergent, and Neglected Viruses Surveillance Laboratory (ViVER) at Fiocruz, Amazonas, Brazil (8). Library preparation was carried out with Illumina’s COVIDSeq kit, and sequencing was carried out on the MiniSeq version 2.3.0 sequencer with the Mid-Output Reagent Cartridge kit (300 cycles) for a 151 bp x 2 pair-end run. FASTQ reads were generated at Illumina BaseSpace (https://basespace.illumina.com) and imported into Geneious Prime v2024.0.5. Initially, reads were trimmed for quality, duplicates were removed, and the remaining reads were normalized and assembled into contigs using a custom workflow that employs the tools BBDuk, Dedupe, BBNorm, and BBMap (v.39.06), embedded into Geneious Prime. The sequences AF484424, AF441119, and AY237111 from Genbank were used as reference sequences for the S, M, and L virus segments of OROV, respectively. The consensus sequences were extracted using a threshold of at least 50% to call a base.

### OROV whole-genome genotyping

The 34 complete OROV sequences of L, M, and S genomic segments generated here were aligned with corresponding segments of a reduced dataset containing the prototype sequences of OROV (L: AF484424; M: AF441119; S: AY237111), Iquitos virus (L: KF697142; M: KF697143; S: KF697144), Perdões virus (KP691627; M: KP691628; S: KP691629), and Madre de Dios virus (L: KF697147; M: KF697145; S: KF697146), as well as OROV sequences representing the major M (M1 and M2), L (L1 and L2) and S (S1, S2 and S3) clades described by Naveca et al., 2024 (8). Sequences were aligned with MAFFT v7.490 (9), and maximum likelihood (ML) phylogenetic trees were inferred with IQ-TREE v2.1.1 (10) using the best substitution model defined by ModelFinder (11) Branch support was inferred using ultra-fast bootstrap (10) on 1,000 replicates. The ML trees were visualized and edited using FigTree v.1.4.4 (https://github.com/rambaut/figtree/releases).

### Bayesian evolutionary and phylogeographic analyses

To reconstruct the spatiotemporal history of OROV spread in Bolivia, we select a subset of near-full-length sequences of the OROV_BR-2015-2024_ epidemic clade that includes the most basal sequence sampled in the Amazonas state in 2015, plus all sequences belonging to the sub-clades AMACRO-I and AMACRO-II (which comprises all sequences sampled in Bolivia, as observed in the ML inference). The resulting dataset was submitted to ML phylogenetic reconstruction as described above, and the temporal structure was estimated by performing a root-to-tip linear regression with TempEst v.1.5.3 (12), in which the significance of the correlation between the collection date and genetic divergence was accessed with a Spearman correlation test. Time-scaled phylogeographic trees for concatenated genomic segments were estimated using the Bayesian Markov chain Monte Carlo (MCMC) approach, implemented in the software BEAST v1.10 (13). Time-scaled trees were inferred with a relaxed uncorrelated lognormal distributed molecular clock model with a continuous-time Markov chain (CTMC) rate reference prior (14) and the non-parametric Bayesian skyline coalescent demographic model (15). The ancestral node states were reconstructed with a CTMC prior, and a discrete spatial diffusion with a symmetric substitution model complemented with a Bayesian stochastic search variable selection (BSSVS) to identify the significant migration routes (16). Markov Chain Monte Carlo (MCMC) was run for 100 million generations, and convergence was assessed by calculating the Effective Sample Size (ESS) for all parameters using Tracer v1.7.1 (17). The maximum clade credibility (MCC) trees were summarized with TreeAnnotator v.1.10 and visualized using FigTree v.1.4.4.

### Statistical analyses

The Chi-square test was used to compare the frequency of symptoms between males and females infected with OROV (two-tailed) and the distribution of OROV cases by age among males and females. The threshold for statistical significance was set to *P* < 0.05. Statistical analyses were performed using GraphPad Prism version 9.0 software.

### Data availability

All OROV consensus sequences generated in this study (n = 102, corresponding to the three segments of 34 unique samples) were deposited in GenBank (https://www.ncbi.nml.nih.gov/genbank) under the accession numbers PQ634468 - PQ634569.

### Ethics statement

This study was approved by the Ethics Research Committee of the Universidad Cristiana de Bolivia (FWA00028929) under protocol number CEI/001/2024.

## RESULTS

### Epidemiological data of Oropouche fever cases in Bolivia

As of EW 35 of 2024, 356 Oropouche cases were confirmed by real-time RT-PCR in Bolivia (**Figure 1a**). The first OROV-positive cases were reported in samples collected from the Pando department during EW 3 of 2024, and the peak of cases occurred in EW 15, coinciding with the Amazon region’s rainy season (from November to April). Oropouche cases have decreased since EW 16, with the last cases reported in EW 20. The majority of cases were identified in the La Paz department (75%), followed by Beni (21%) and Pando (3%) (**Figure 1b**). The cases were reported among 16 municipalities, with the highest proportion of cases reported in the municipalities of Irupana (33%), La Asunta (13%), and Chulumani (12%) in the La Paz department, and in the municipality of Guayaramerín (12%) in the Beni department. Oropouche cases were evenly distributed between males and females, but there was a significant difference in the distribution of infections by age group between the sexes (*P* = 0.0496). The highest proportion of Oropouche cases occurred in the 30-39 age group for females (25%) and in the 10-19 age group for males (25%) (**Figure 1c**). The most common symptoms reported for OROV infections were fever (97%), headache (94%) and myalgia (82%), followed by nausea (41%), vomiting (27%), loss of appetite (12%) and photophobia (10%) (**Figure 1d**). Notably, nausea and vomiting were more frequently observed in females (46% and 39%, respectively) than in males (34% and 13%, respectively), although the difference was only significant for vomiting (*P* = 0.0006). All OROV cases were mild, and no severe cases involving the central nervous system or deaths associated with OROV were reported.

**Figure 1.**
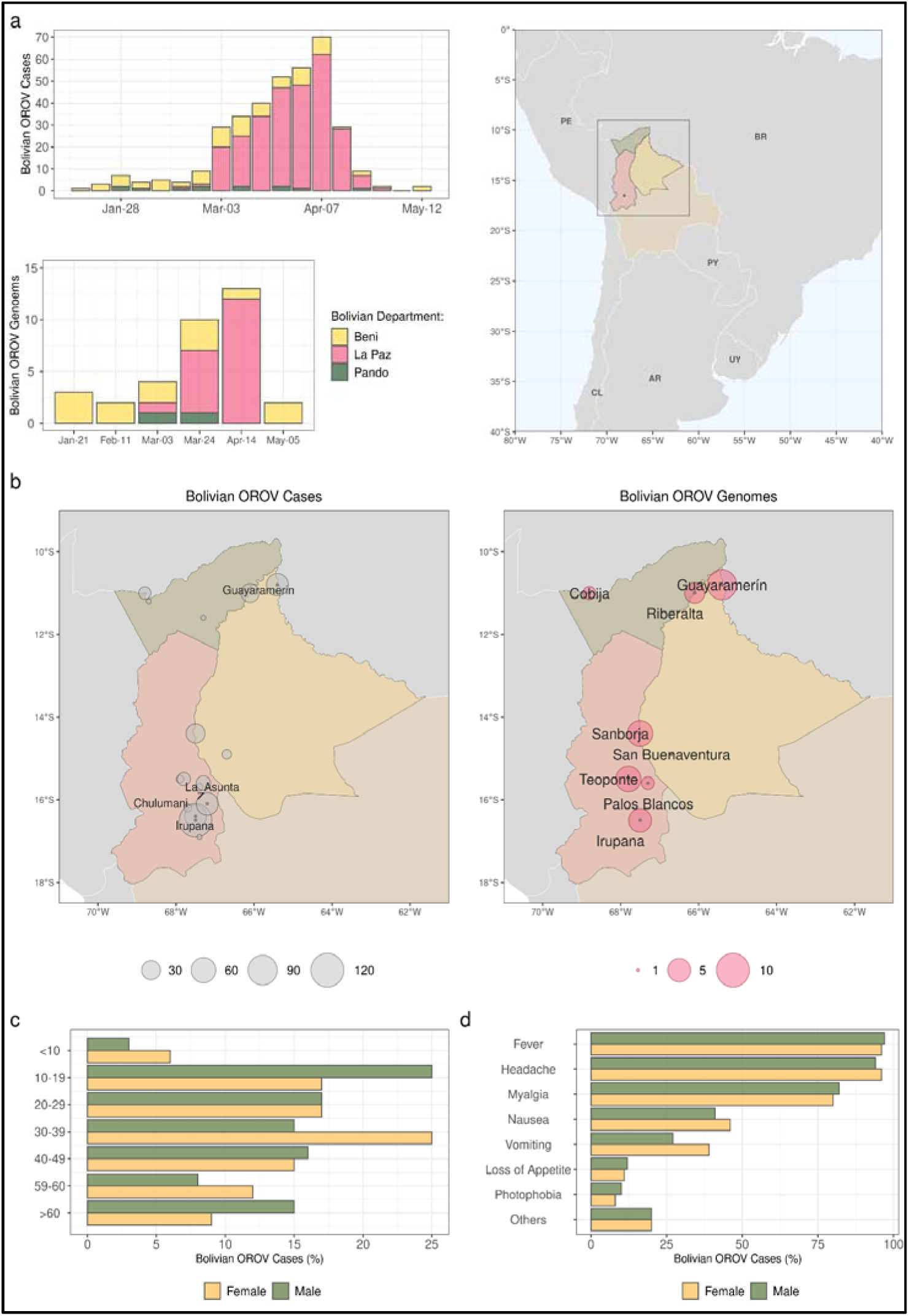
The 2024 Oropouche virus outbreak in Bolivia. **a)** Temporal distribution of Oropouche cases and sampled genomes across Bolivian departments, with the corresponding map on the panel’s right side. The graphs and the map are color-coded according to the legend at the bottom of the panel. **b)** Spatial distribution of Oropouche cases and sampled genomes across Bolivian cities. In both maps, the circle’s diameter is proportional to the number of cases and genomes, as indicated by the respective legends. Only cities with more than five genomes are labeled in the first map, while in the second map, all cities with sampled genomes are annotated. **c)** Proportion of OROV-positive cases in Bolivia, categorized by age and gender. **d)** The proportion of symptoms recorded from OROV-positive cases in Bolivia is stratified by gender. AR: Argentina, BR: Brazil, CL: Chile, PE: Peru, UY: Uruguay.

### Spatiotemporal patterns of the OROV spread across Bolivia

A total of 34 full-length OROV genomes were generated from samples collected between EW3 and EW20 from all three Bolivian departments (Figure 1b). The ML phylogenetic analyses performed individually for each genomic segment revealed that all 34 OROV sequences belong to the novel reassortant OROV_BR-2015-2024_ clade that drove the recent epidemic in Brazil (8) (Figure 2). The ML phylogenetic analysis of the OROV_BR-2015-2024_ clade performed with concatenated segments L, M, and S indicates that OROV sequences from Bolivia are nested within the Brazilian sub-clade AMACRO-II, which is one of the major sub-clades identified in the Brazilian Amazonian region (Figure 3). The sub-clade AMACRO-II most probably emerged in the Acre state around August 2023 and comprised all OROV sequences detected in the Brazilian states of Acre and Rondônia, which borders Bolivia, and all sequences from the southern region of the Amazonas state sampled from December 2023 onwards (8).

**Figure 2.**
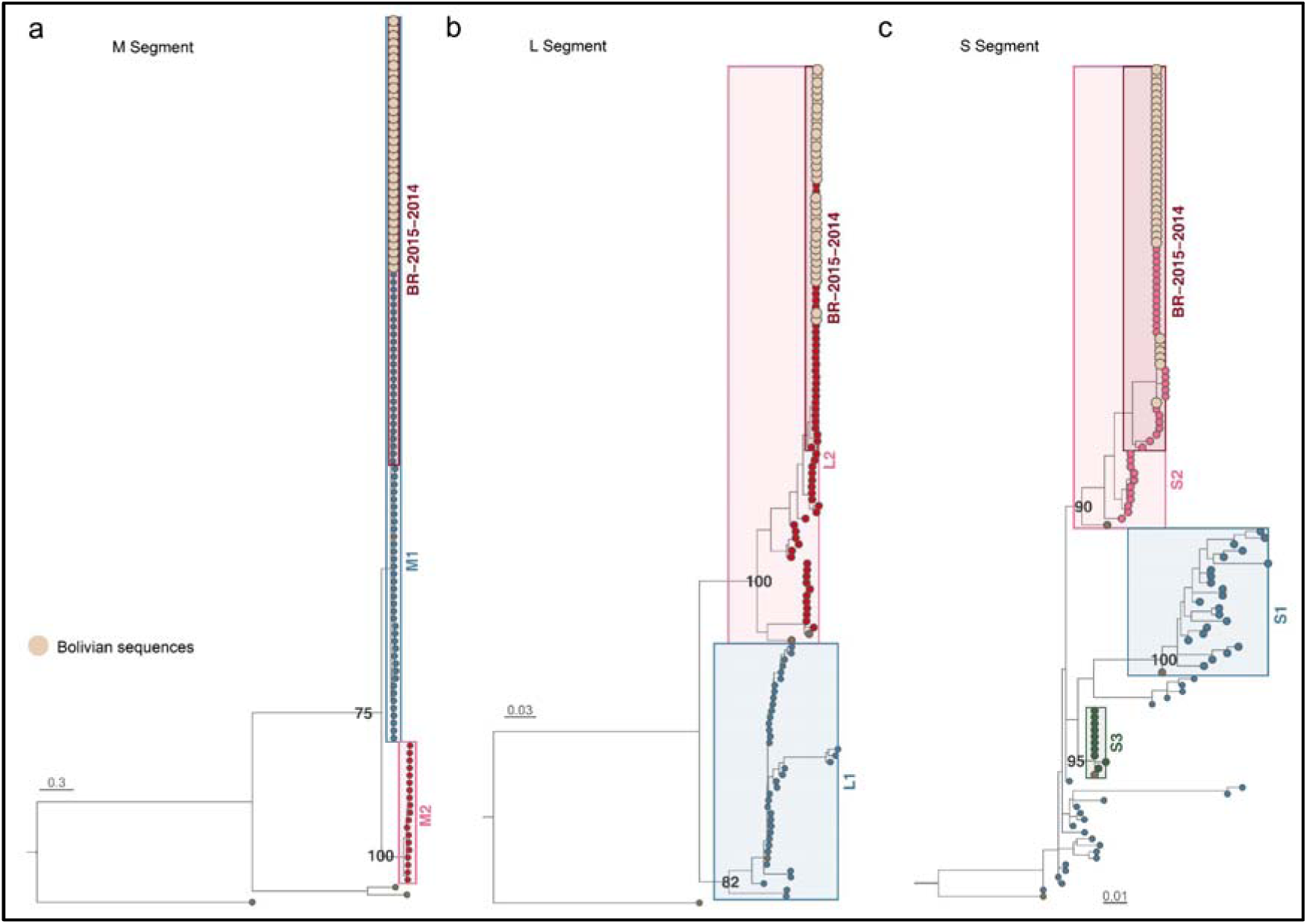
ML phylogenetic analyses of the M, L and S segments of OROV. Phylogenetic trees inferred from the segments M (a), L (b), and S (c) of OROV sequences with complete genomes (n = 131). The tips of Bolivian viruses are colored in light brown with an increased diameter, and prototypical OROV sequences in darker brown. Brackets demarcate major OROV clades alongside their denomination and statistical support (aLRT). All trees are drawn according to the genetic distance scale at the bottom of each panel.

**Figure 3.**
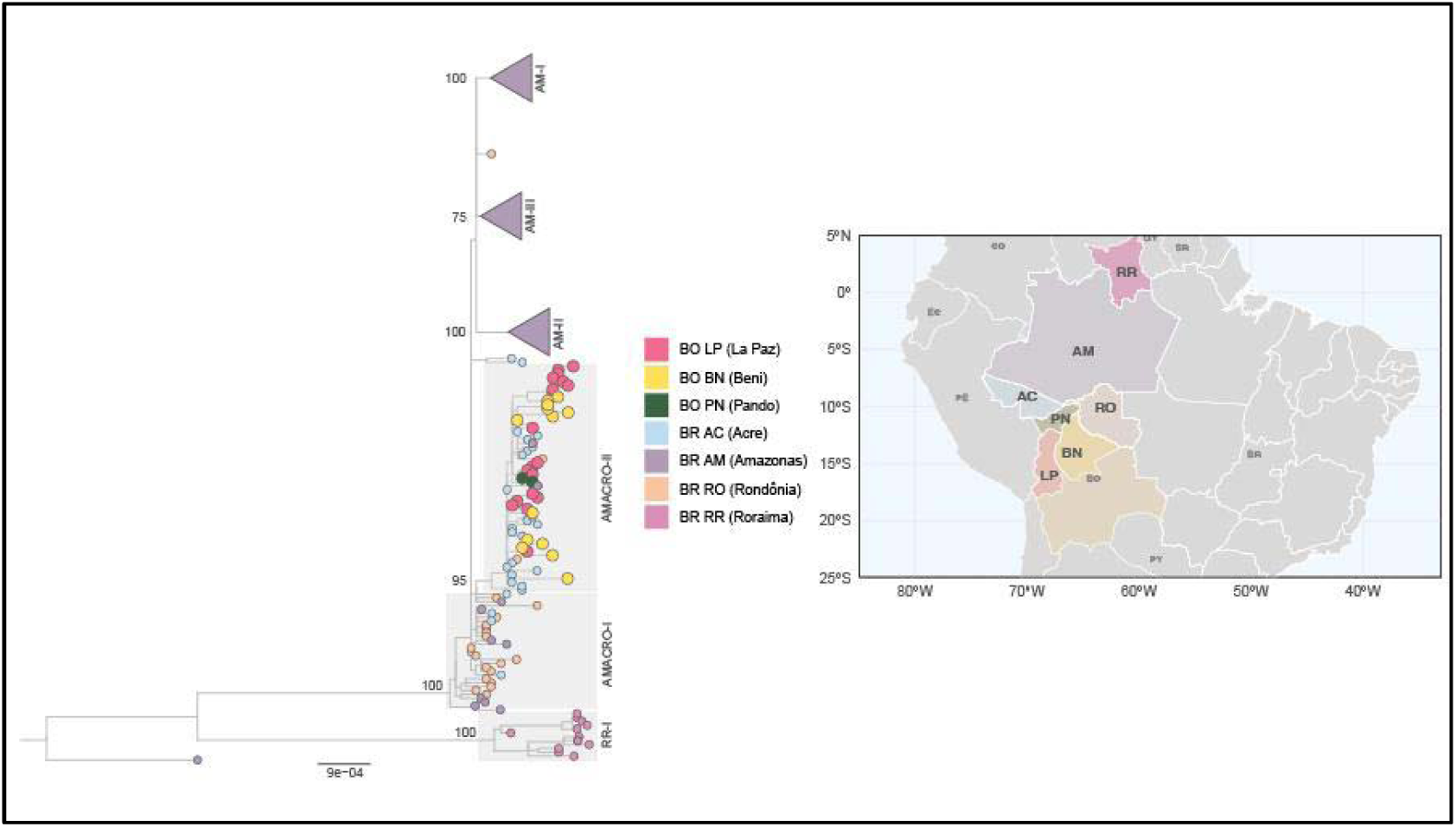
ML phylogenetic tree of concatenated segments of sequences belonging to the OROV_BR-2015-2024_ clade. Major OROV_BR-20125-2024_ sub-clades are highlighted with their respective designations and annotated statistical support (a*LRT*). Tip points are color-coded according to their sampling locations, which are also reflected on the map displayed on the left side of the panel. Bolivian sequences are marked with slightly larger shapes for easier identification. To improve visualization, the sub-clades from the Brazilian state of Amazonas are collapsed into triangles. The tree is drawn according to the scale at the bottom of the left panel. AM: Amazonas, BO: Bolivia, BE: Beni, BR: Brazil, CO: Colômbia, EC: Ecuador, LP: La Paz, PE: Peru, PN: Pando, PY: Paraguay, RO: Rondônia, RR: Roraima.

Bayesian phylogeographic analysis with concatenated L, M, and S segments was then performed to model the viral diffusion process between Brazil and Bolivia in a discrete space. For this analysis, we retained the oldest sequence of OROV_BR-2015-2024_ clade detected in the Amazonas state in 2015, and all OROV sequences belonging to sub-clades AMACRO-I (detected between January and June 2023) and AMACRO-II (detected from December 2023 onwards). OROV sequences from Bolivia, Acre, Amazonas, and Rondônia were grouped by municipality of origin. None of the patients in the present study reported having a travel history. The correlation between genetic divergence and sampling time was significant for this OROV dataset (p < 0.05) (**Figure 4a**), supporting a robust temporal structure. The genome-wide molecular clock rate of the dataset was estimated at 1.2 x 10^-3^ (95% Highest Posterior Density [95% HPD]: 1.0 - 1.5 x 10^-3^) substitutions/site/year, similar to that previously estimated for the complete dataset of OROV_BR-2015-2024_ clade (8).

**Figure 4.**
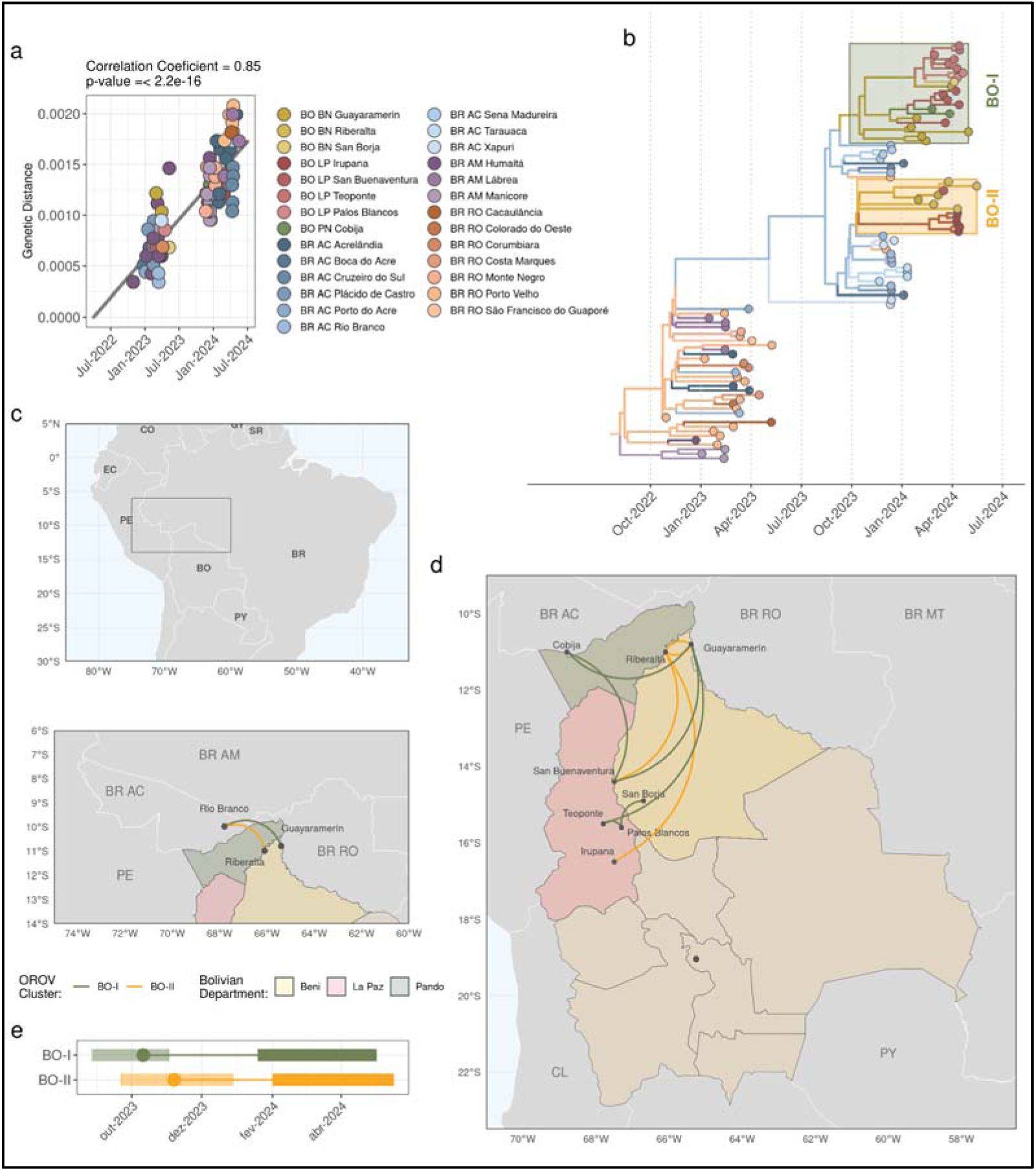
Spatio-temporal dynamics of the OROV spread in Bolivia. **a.** Linear regression of root-to-tip genetic divergence against sampling dates for sequences (*n* = 92) in the OROV_BR-2015-2024_ subset. Samples are color-coded by sampling city, as per the color scheme shown on the right side of the panel. **b.** Time-scaled MCC tree of the OROV_BR-2015-2024_ sequences circulating in Bolivia and neighboring Brazilian states of Acre, Rondônia, and the southern region of Amazonas. Tips are color-coded by sampling location, and branches are colored according to inferred ancestral locations, following the scheme on the previous panel. Bolivian clusters BO-I and BO-II are annotated in the tree. **c.** Map of Bolivia and its neighboring countries. The selected region, which encompasses parts of northern Brazil and the northernmost area of Bolivia, is shown in greater detail at the bottom of the panel. The lines indicate the introductions that established the subclusters BO-I and BO-II, connecting their origins in Brazil to destination cities in Bolivia. Migrations are color-coded according to the Bolivian cluster, as indicated in the legend at the bottom of the panel. **d.** Map of Bolivia highlighting the sub-national departments. Lines represent inferred viral migrations as reconstructed through discrete phylogeographic analysis, connecting origins and destinations of migrations with arcs oriented clockwise and color-coded consistently with the previous panel. **e.** The median T_MRCA_ (circle) and its 95% HPD interval (transparent polygon) of the BO-I and BO-II subclusters are shown, along with the period of cryptic circulation (thin line) and the sampling range (thick line). AC: Acre, AM: Amazonas, BN: Beni, BR: Brazil, CL: Chile, CO: Colômbia, EC: Ecuador, RO: Rondônia, LP: La Paz, PE: Peru, PN: Pando, PY: Paraguay.

Bayesian phylogeographic analysis revealed that the 34 OROV sequences from Bolivia formed two monophyletic sub-clades, designated BO-I (*n* = 22) and BO-II (*n* = 12) (**Figure 4b**). Rio Branco, the capital of Acre state, was identified as the most probable source of the Bolivian sub-clades BO-I (posterior state probability [*PSP*] = 0.79) and BO-II (*PSP* = 1) (**Figure 4c**). The BO-I and BO-II sub-clades were likely introduced into the nearby municipalities of Guayaramerín (*PSP* = 0.92) on October 12, 2023 (95% HPD: September 5 – November 17, 2023) and Riberalta (*PSP* = 0.84) on November 10, 2023 (95% HPD: September 24 – December 31, 2023), respectively, both located in the Beni department (**Figure 4c**). From Guayaramerín, the BO-I sub-clade spread westward to the municipality of Cobija (Pando department) and southward to Teoponte (La Paz department) (**Figure 4d**). The BO-I sub-clade further spread from Cobija to San Buenaventura (La Paz department), from Teoponte to Palos Blancos (La Paz department), and from Palos Blancos to San Borja (Beni department) (**Figure 4d**). From Riberalta, the BO-II sub-clade spread eastward to the municipality of Guayaramerín and southward to San Buenaventura and Irupana (La Paz department) (**Figure 4d**). The lag between the median time to the most recent common ancestor (T_MRCA_) and the earliest genome sequence for each Bolivian sub-clade was 99 days for BO-I and 82 days for BO-II (**Figure 4c**).

## DISCUSSION

In 2024, Bolivia experienced its largest documented Oropouche fever outbreak to date, affecting 16 municipalities in three departments of the Amazon basin (Pando, Beni, and La Paz). Interestingly, during this outbreak, no cases of Oropouche were reported in the department of Cochabamba, where the first cases of Oropouche were reported between 2005 and 2007 (3). The 2024 OROV epidemic in Bolivia spanned from January to May and peaked in mid-April, thus coinciding with the rainy season (November to April) (https://www.ine.gob.bo/index.php/medio-ambiente/clima-y-atmosfera/). The same phenomenon was observed in the Brazilian Amazon Region, where the increase in OROV circulation coincides with the rainy season (December to March) (8). This is likely linked to the expansion of humid habitats, which create favorable conditions for the development of *C. paraensis* larvae (18,19). Further studies are required to confirm the correlation between the onset of the rainy season, the increased presence of midges, and the occurrence of OROV outbreaks in Bolivia’s Amazon region.

Most Oropouche cases during the 2024 epidemic in Bolivia were mild, and the most common clinical manifestations (fever, headache, and myalgia) were the same as those observed in previous (20) and current (8,21) OROV epidemics. A key difference was the very low incidence of arthralgia (<2%) and the complete absence of retro-orbital pain in Bolivia, both of which were reported at much higher frequencies (30-40%) (8,20). Notably, we detect a higher frequency of nausea and a significantly higher frequency of vomiting in females (46% and 39%, respectively) than in males (34% and 13%, respectively) in Bolivia, a finding entirely consistent with a recent study conducted in the Amazonas state (8). Since nausea and vomiting may be signs of liver disease, these findings could indicate a greater likelihood of liver damage in females compared to males. We also identified gender differences in the distribution of OROV cases across age groups, potentially indicating higher exposure to the vector at younger ages among males compared to females.

Phylogenetic analyses confirm that all OROV cases detected in Bolivia belong to the OROV_BR-2015-2024_ clade, which originated from a new reassortment event around 2010-2015 and that was responsible for the increase of OROV cases in the Brazilian western Amazon region between 2022 and 2024 (8). These analyses further revealed that the OROV sequences from Bolivia were part of the AMACRO-II sub-clade, which circulates in the Brazilian states of Acre and Rondônia since late 2023 (8), two states that border the departments of Pando and Beni, respectively, where the first Oropouche cases were detected in Bolivia. The discrete phylogeographic analysis indicates that the AMACRO-II sub-clade spread from Rio Branco, the capital of Acre state, to the municipalities of Guayaramerín and Riberalta in the Beni department, which borders the Brazilian state of Rondônia. Thus, the OROV AMACRO-II sub-clade was repeatedly transmitted across the neighboring Amazonian regions of Acre, Rondônia, and Beni in the bordering region between Brazil and Bolivia.

Our temporal reconstruction supports that the BO-I subclade was probably introduced in Guayaramerín around October 2023, and the BO-II subclade in Riberalta around November 2023. Therefore, we estimated that OROV circulated cryptically between two and three months before its first detection in Bolivia in January 2024. Both Bolivian OROV subclades were successfully disseminated beyond the Beni department to the departments of Pando (BO-I) and La Paz (BO-I and BO-II), all located in the Amazon basin. This confirms OROV’s capacity for rapid spread throughout the Bolivian Amazon region, which is approximately 65% of Bolivia’s territory. Importantly, OROV is also currently spreading outside the Brazilian Amazon basin (22,23), which suggests the importance of strengthening the differential diagnosis of OROV and intensifying entomological surveillance for the OROV vector in Bolivian departments beyond the Amazon basin, like south of La Paz, Potosí, Oruro, south of Cochabamba and Tarija.

The precise causes of the recent upsurge of OROV in Bolivia are unclear. First, estimating the true prevalence of OROV infection in Bolivia during previous decades is challenging, as systematic surveillance for the virus was not in place before 2024. The recent surge in OROV cases may be associated with the spread of a novel reassortant OROV lineage. However, the relative transmissibility of this new lineage compared to those circulating in previous years has not yet been evaluated. Moreover, the recent spread of a novel reassortant OROV in the Amazon region coincided with a remarkable El Niño Southern Oscillation (ENSO) event, which caused above-average temperatures (maximum and minimum) in all Bolivian departments in the period from November 2023 to March 2024 (24). This climatological factor, along with agricultural expansion and deforestation, may have contributed to the transmission of OROV in Bolivia (25,26).

The major limitation of our study is the geographic-based sampling bias due to the low number or absence of sequence data from some key locations in Bolivia and neighboring Brazilian regions. The two most probable entrance points of OROV in Bolivia, Guayaramerín and Riberalta, are locations closer to Rondônia state than Acre state. Notably, the Bolivian city of Guayaramerín is located on the left bank of the Mamoré River, in front of the Rondônia city of Guajará-Mirim, in Brazil. These twin cities display an intense and dynamic daily trade flow (27), potentially making them a critical area for the transborder spread of OROV. Most Brazilian sequences in the AMACRO-II sub-clade sample so far, however, were collected in the Acre state. Despite experiencing a significant OROV epidemic in 2024, Rondônia has only a limited number of viral sequences from this year described so far, and this geographic-based sampling bias may have impacted the accuracy of discrete phylogeographic inferences presented here.

In conclusion, our study confirms that the novel Brazilian OROV_BR-2015-2024_ clade was introduced at least two times and cryptically circulated for 2-3 months in Bolivia. The spatiotemporal pattern supports that OROV dissemination was initially driven by short-distance movements from the Brazilian states of Acre or Rondônia to the neighboring Bolivian department of Beni and subsequent spread from Beni to other Bolivian departments. These findings demonstrate the epidemic potential of the new reassortant OROV lineage to spread beyond Brazil’s borders and underscore the importance of maintaining active surveillance for emerging and reemerging viruses, particularly in cross-border regions of the Amazon shared by Bolivia and Brazil.

## Acknowledgements

We want to thank to the Pan American Health Organization, Washington D.C., USA, (Infectious Hazards Management Unit, Health Emergencies Department) for their continued support in providing reagents for sequencing; The General of Directorate of Epidemiology from the Bolivian Ministry of Health for their support this study providing epidemiological data; Oswaldo Cruz Institute, Fiocruz, Rio de Janeiro, RJ, Brazil (Laboratory of Arboviruses and Hemorrhagic Viruses) and Leônidas and Maria Deane Institute, Fiocruz, Manaus, AM, Brazil (Center for Surveillance of Emerging, Reemerging or Neglected Viruses – ViVER/EDTA) for their support in the analysis of sequencing data and writing of this article.

## Authors’ contribution

JACV, SSM, COM and LRLM did the genomic sequencing. JACV, IA, FGN, SSM, GB and LRLM performed the bioinformatic work and phylogenetic analyses. JACV, GB, IA, SSM, LG, MMG, FGN and LRLM wrote the article. HCL, RSK and MXEM assisted with the epidemiological analysis. JACV, IA, SSM, COM, NML, JDM, LG, MMG, JMR, GB, FGN and LRLM contributed to the overall design, reviewed, and commented on draft articles.

---

**Table S1.** Epidemiological information of the OROV positive sequenced samples.

| Sample ID | EW | Departament | Municipality | Age range | Sex | Hospitalization |
| --- | --- | --- | --- | --- | --- | --- |
| 3960-24 | 3 | BENI | Guayaramerín | 16-20 | Male | Unknown |
| 3969-24 | 4 | BENI | Guayaramerín | 31-35 | Female | No |
| 3982-24 | 5 | BENI | Guayaramerín | 11-15 | Female | No |
| 3801-24 | 6 | BENI | Guayaramerín | 41-45 | Male | Unknown |
| 442-24 | 7 | BENI | Guayaramerín | 41-45 | Male | No |
| 5058-24 | 8 | LA PAZ | Teoponte | 31-35 | Male | Unknown |
| 6147-24 | 8 | PANDO | Cobija | 66-70 | Female | No |
| 4546-24 | 9 | BENI | Riberalta | 21-25 | Male | Unknown |
| 5059-24 | 9 | LA PAZ | Teoponte | 51-55 | Female | Unknown |
| 3730-24 | 10 | BENI | Guayaramerín | 21-25 | Female | Unknown |
| 4808-24 | 11 | LA PAZ | San Buenaventura | 36-40 | Female | No |
| 4709-24 | 12 | BENI | Riberalta | 26-30 | Female | Unknown |
| 4814-24 | 12 | LA PAZ | San Buenaventura | 16-20 | Female | Unknown |
| 4827-24 | 12 | LA PAZ | San Buenaventura | 31-35 | Male | No |
| 4831-24 | 13 | LA PAZ | San Buenaventura | 21-25 | Female | Unknown |
| 5115-24 | 13 | BENI | Guayaramerín | 36-40 | Female | Unknown |
| 5194-24 | 13 | PANDO | Cobija | 16-20 | Female | No |
| 5229-24 | 14 | BENI | San Borja | 11-15 | Male | No |
| 5342-24 | 14 | LA PAZ | Irupana | 11-15 | Female | No |
| 5564-24 | 14 | LA PAZ | Teoponte | 51-55 | Female | No |
| 5239-24 | 15 | LA PAZ | San Buenaventura | Unknown | Male | Unknown |
| 5305-24 | 15 | BENI | Riberalta | 31-35 | Male | No |
| 5327-24 | 15 | LA PAZ | Irupana | 56-60 | Female | Unknown |
| 5339-24 | 15 | LA PAZ | Irupana | 26-30 | Female | Unknown |
| 5343-24 | 15 | LA PAZ | Irupana | 36-40 | Female | Unknown |
| 5389-24 | 15 | LA PAZ | Palos Blancos | 21-25 | Female | No |
| 5555-24 | 15 | LA PAZ | Teoponte | 31-35 | Female | No |
| 5570-24 | 15 | LA PAZ | San Buenaventura | 36-40 | Female | Unknown |
| 5553-24 | 16 | LA PAZ | Teoponte | 51-55 | Male | No |
| 5563-24 | 16 | LA PAZ | Teoponte | 61-65 | Male | No |
| 5686-24 | 16 | LA PAZ | Irupana | 0-5 | Female | Unknown |
| 5825-24 | 17 | LA PAZ | Palos Blancos | 11-15 | Female | No |
| 6094-24 | 18 | BENI | Guayaramerín | 11-15 | Male | No |
| 6397-24 | 20 | BENI | Riberalta | 41-45 | Female | No |

Samples ID used in this table corresponds to the sequencing IDs and can only be decoded to the original samples by the authors of the current study.

